# Impaired ICOS signaling between Tfh and B cells distinguishes hospitalized from ambulatory CoViD-19 patients

**DOI:** 10.1101/2020.12.16.20248343

**Authors:** Amanda Hanson, Heather Cohen, Hao Wang, Nandini Shekhar, Chinmayee Shah, Abha Dhaneshwar, Bethany W. Harvey, Richard Murray, Christopher J. Harvey

## Abstract

Emerging evidence suggests that SARS-CoV-2 infections are characterized by systemic immune responses that appear to be dysregulated with more severe CoViD-19 disease. Lymphopenia and delayed antibody responses are commonly identified in CoViD-19 subjects, and recent reports have demonstrated abrogation of germinal centers in severe CoViD-19. This work assessed a potential mechanistic basis for impaired humoral responses, focusing on the T follicular helper (Tfh) and B cell interface that is critical for germinal center reactions. Here we demonstrated that Tfh activity is impaired in hospitalized relative to ambulatory CoViD-19 subjects, potentially due to decreased expression of the costimulatory molecule ICOS-L on B cells. Functional impairment manifested as a diminished ability to stimulated Tfh derived IFNγ and IL-21, the latter of which is critical for B cell proliferation and differentiation. Activation of Tfh cells by agonism of the ICOS receptor ex vivo by an agonistic antibody stimulated the generation of IFNγ/IL-21 double positive cells from hospitalized CoViD-19 subjects. This report establishes an immunological defect that differentiates ambulatory from hospitalized CoViD and suggests that agents that could restore impaired mechanisms at the Tfh–B cell interface may be of therapeutic value.

## Introduction

The emergence of the SARS-CoV-2 virus from zoonotic transmission and the ensuing CoViD-19 pandemic has resulted in approximately 61.3 million cases and over 1.44 million deaths globally as of November 27, 2020 ^1^. Aberrant activation or dysfunction of the immune system as it relates to progressive CoViD-19 disease has been the focus of recent investigations ^2, 3, 4, 5, 6, 7, 8, 9^. The adaptive immune response is driven by the activity of T and B lymphocytes, and a consistently observed immunological feature of progressive disease has been a negative association of both T and B cell frequencies with increasing CoViD-19 severity ^2, 3, 4, 5, 7, 9^. This reduction in lymphocyte populations may contribute to a protracted course of disease and the delayed or absent humoral immune response observed in severe cases.

Assessment of T cell responses have demonstrated a progressive decline in overall CD3+ T cells, and consensus across studies points to the decline occurring in both CD4 and CD8 T cell populations ^3^. This decrease is consistently observed in a number of infectious diseases, including pandemic influenza; however, the duration of lymphopenia is significantly longer in CoViD-19 ^10, 11^. Despite the decreasing frequencies and prolonged lymphopenic state of CoViD-19 subjects, generation of antigen-specific T cell responses appears to be intact as viral CD4 and CD8 responses have been detected in virtually all convalescent subjects ^6, 8^.

While memory T cells maintain their activity, initiation of B-cell mediated immune responses appears to be delayed and also short-lived ^9, 12, 13, 14^. CD4 Tfh cells are the critical cell type for initiating germinal center reactions and stimulating the humoral response to viral pathogens. Limited data to date have suggested that Tfh cells are increased within the CD4 compartment in CoViD-19 subjects ^3, 7, 9^. The paradoxical increase in Tfh cells coupled with B cell lymphopenia and delayed antibody responses suggest impairment of germinal center responses. The absence of intact germinal centers in severe CoViD-19 cases has been recently demonstrated in studies where pathological assessment of hilar lymph nodes in autopsied subjects as well as in lymph node and spleen biopsies from infected subjects were performed ^15, 16^. Among the mechanisms critical for germinal center formation, in both animal systems and in humans, it is well established that the ICOS-L / ICOS signaling pathway plays a critical role ^17, 18, 19, 20, 21, 22, 23^. In this study, we set forth to examine peripheral blood Tfh and B cells in samples from human CoViD-19 donors of various disease severity to elucidate functional capacity and identify mechanisms of any observed defect that may be contributing to the discordant observations of Tfh activation but impaired GC activity in CoViD-19, with the ultimate goal of identifying actionable therapeutic targets.

## Results

### Peripheral Blood Phenotyping Suggests Altered Tfh/B Cell Axis and Impaired Co-Stimulation

Three cohorts were used as part of this study and represented CoViD-19-naïve donors (Healthy Donors), ambulatory CoViD-19 donors (Ambulatory), and hospitalized CoiViD-19 donors (Hospitalized). Detailed description of study cohorts is provided in the supplemental material and in Table S1. Flow cytometry-based immunophenotyping of samples from all three cohorts was performed to assess the frequencies of immune cell populations within the peripheral blood compartment compared to published findings. Consistent with the literature, flow profiling demonstrated a reduction in CD19+ B cells, with decreases observed in both Ambulatory (mean frequency of 6.2% of leukocytes) and Hospitalized cohorts (mean frequency of 8.8% of leukocytes) relative to healthy donors (phenotyping cohort; mean frequency of 12.6% of total leukocytes) (Fig. 1A). No significant changes were observed within CD8 T cells (CD3+ CD19-CD4-) (Fig. 1C), however a significant decrease in CD4 T cells (from mean frequency of 32.8% to 17.7% of total leukocytes, Fig. 1D) and corresponding trend towards an increase in CD3-CD19-monocytes (from mean of 39.2% to 59.6%, Fig. 1B) were only observed in the Hospitalized CoViD-19 cohort.

**Fig. 1:**
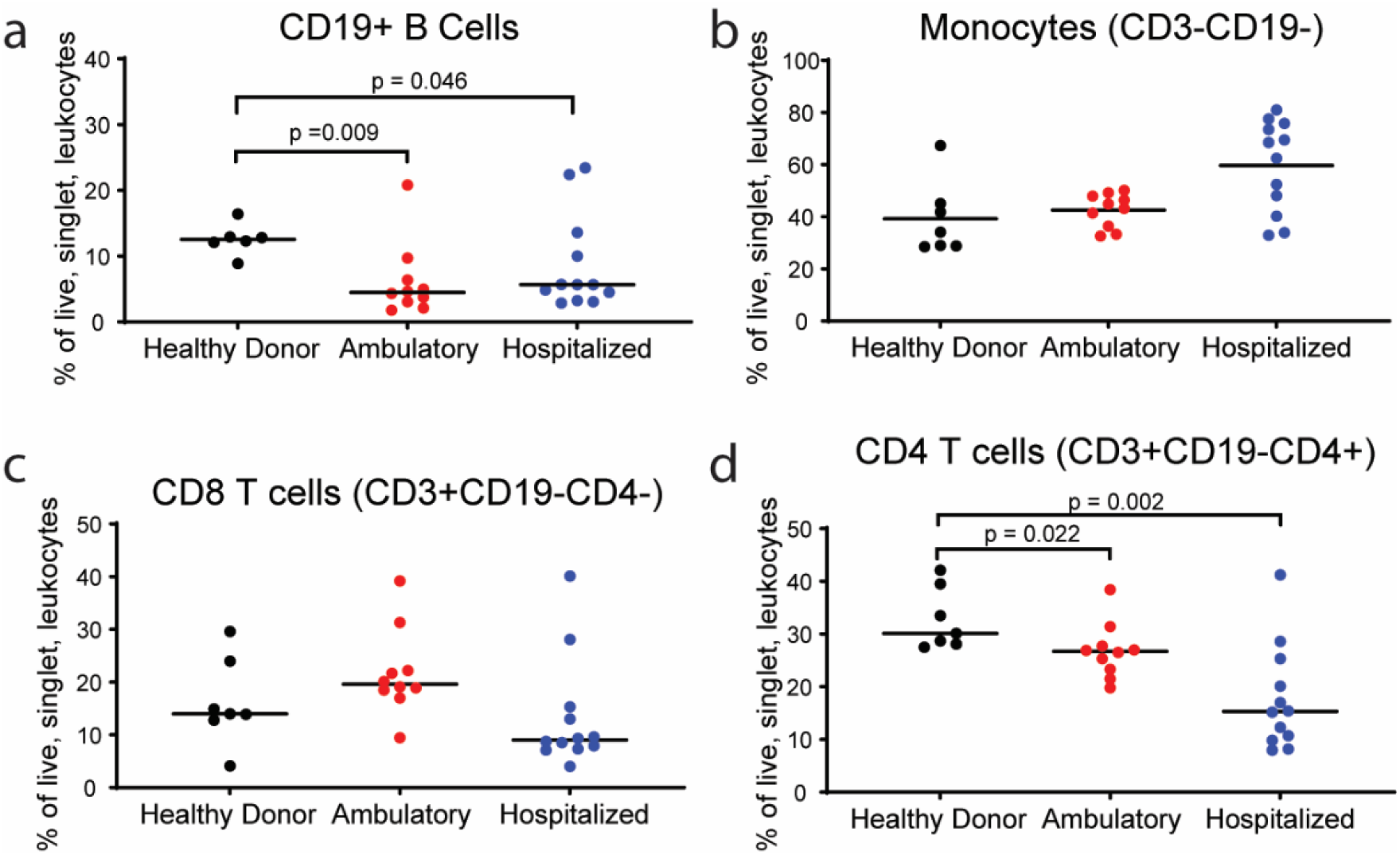
CD4+ T cells decline with disease severity in CoViD-19 subjects. **a-d**, The comparison of immune subset frequencies in the blood. a, CD19+ B cells of healthy donors (phenotyping cohort; n=6), Ambulatory (n= 10) and Hospitalized (n=12) CoViD-19 donors; b, myeloid cells; c, CD4-T cells; d, CD4+ T cells of healthy donors (n=7), Ambulatory (n= 10) and Hospitalized (n=12) CoViD-19 donors. Means are shown (black line).

T cell and B cell populations are interdependent on a variety of levels, and Tfh cells, specifically, are critical mediators of antiviral B cell responses ^18, 21, 22, 23^. As established in influenza infection and vaccination, circulating Tfh populations are an induced, transient, and antigen specific population of CD4 T cells whose presence correlates to long-term antibody responses and protection from either re-infection or infection following vaccination. To assess whether circulating Tfh (cTfh) cell populations are changing within CoViD-19 subjects, phenotypic profiling, including assessment of activation status was performed on peripheral blood mononuclear cells (PBMCs). This analysis demonstrated a significant enrichment of cTfh cells (defined as CD3+ CD4+ CXCR5+ ICOS+ lymphocytes) within Ambulatory subjects and encompassed on average 15% of the peripheral blood CD4 compartment relative to the 8% cTfh population observed in healthy donors (Fig. 2A). This increase, however, was limited to the Ambulatory cohort, and cTfh cells were significantly decreased relative to both healthy donors and Ambulatory CoViD-19 subjects and represented only 2% of CD4 T cells in Hospitalized subjects. To assess whether age may be confounding the observed differences in Tfh frequency, cellular phenotype and frequency were assessed in age matched healthy donor cohorts, and no decrease was observed in age-matched healthy donor subjects (Fig. S3), indicating that the reduction in Hospitalized CoViD-19 subject Tfh cells does not appear to be generally age-related.

**Fig. 2:**
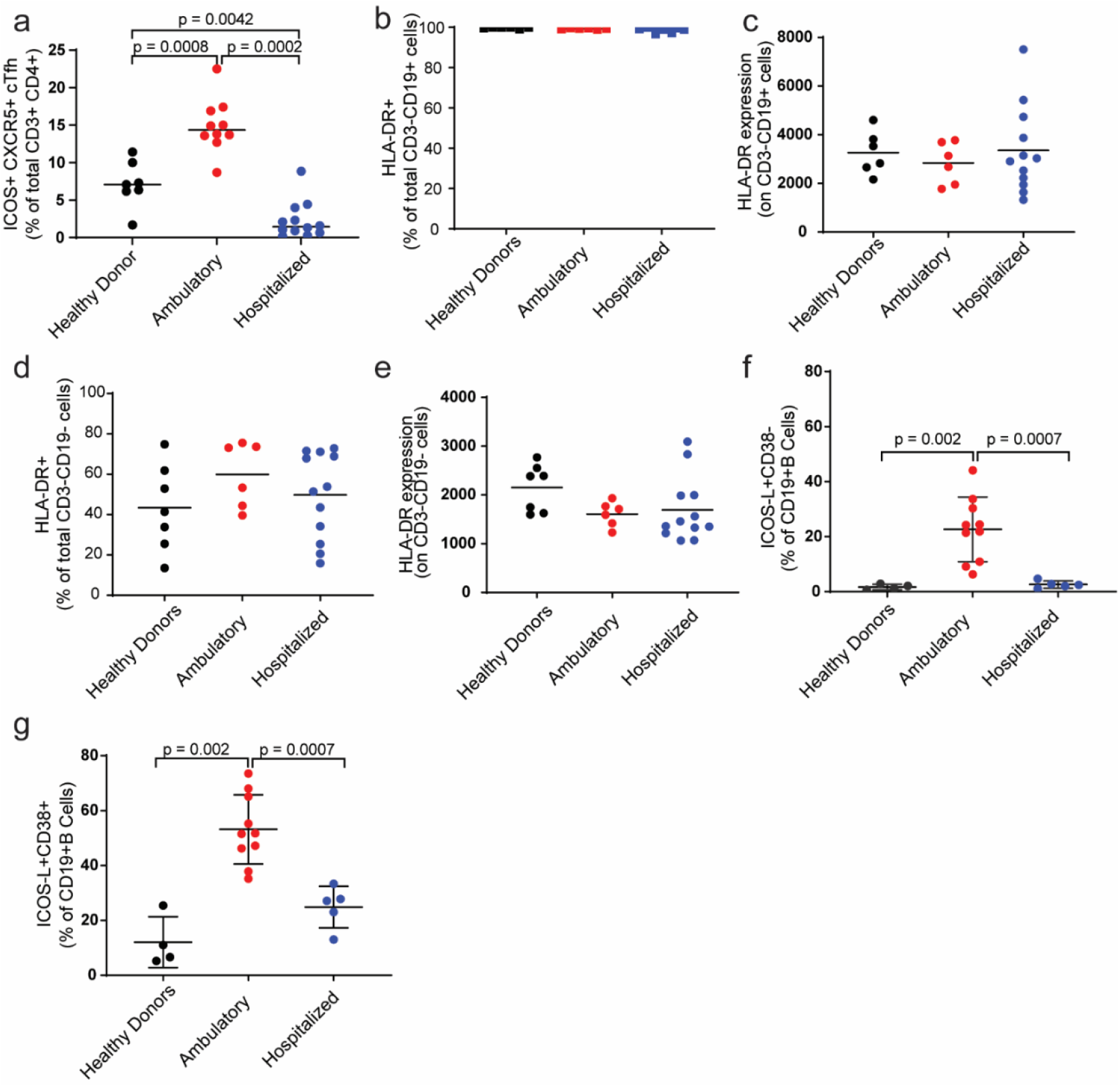
Significant Reduction in cTfh Cells is Observed in Hospitalized CoViD-19 Donors and is Associated with Lack of ICOS-L Mediated Co-stimulation by B cells. **a**, The comparison of cTfh frequencies in the blood of healthy donors (phenotyping cohort; n=7), Ambulatory (n= 10) and Hospitalized (n=12) CoViD-19 donors. **b**, The comparison of HLA-DR+ B cell frequencies in blood and **c**, HLA-DR expression on B cells across donor cohorts in the blood of healthy donors (phenotyping cohort; n=6), Ambulatory (n= 6) and Hospitalized (n=12) CoViD-19 donors. **d**, The comparison of HLA-DR+ myeloid frequencies in blood and **e**, HLA-DR expression on myeloid cells across donor cohorts in the blood of healthy donors (phenotyping cohort; n=7), Ambulatory (n= 6) and Hospitalized (n=12) CoViD-19 donors **f**, The comparison of ICOS-L+ CD38- and **g**, ICOS-L+ CD38+ B cell frequencies in the blood of healthy donors (phenotyping cohort; n=4), Ambulatory (n= 10) and Hospitalized (n=5) CoViD-19 donors. Means are shown (black line).

Within the Hospitalized CoViD-19 donors, the observed decrease in peripheral CD4 T cells and B cell populations may be due to a significant alteration of the Tfh/B cell axis in severe disease. To test this hypothesis, assessment of various mechanisms that may contribute to defective Tfh/B cell activation were interrogated. Tfh-mediated B cell activation is primarily driven by three distinct processes: antigen-presentation by B cells or other APC, B-cell mediated co-stimulation of T cells, and cytokine-driven B cell activation and differentiation. Assessment of phenotypic markers by flow cytometry demonstrated an intact antigen-presentation capacity across cohorts, as there were no observed differences in HLA-DR expression on CD19+ B cells as assessed as either fraction of HLA-DR+ cells (Fig. 2B) or in terms of per-cell expression as assessed by mean fluorescence intensity (Fig. 2C). Similar analysis of HLA-DR expression on monocytes demonstrated trends towards increased HLA-DR expression in CoViD-19+ subjects when viewed as a % of HLA-DR positive cells (Fig. 2D), but expression trended lower on a per-cell basis (Fig. 2E). Together, the data suggest intact antigen presentation capacity within CoViD-19 subjects, and both HLA-DR+ monocytes and B cells appear capable of presenting viral antigens to stimulate initial TCR priming.

To further drive T cell responses, co-stimulation is necessary, as the absence of co-stimulation can drive anergy ^24^. Studies examining the Tfh-B cell entanglement within the germinal centers have demonstrated a critical role for the ICOS/ICOS-L pathway for driving the development of humoral responses ^21^. B cell expressed ICOS-L activates the co-stimulatory receptor ICOS on CD4 Tfh cells, which in turn has been shown to drive a feed-forward mechanism to promote B cell activation and differentiation through a variety of accessory molecules, including CD40L and the cytokine IL-21 ^19, 20, 21, 22, 25^. Profiling of cTfh cells demonstrated increased ICOS+ Tfh cells within Ambulatory cohorts but a significant reduction in more severe subjects. To assess for potential defects in co-stimulation of CD4 T cells by B cells, we profiled ICOS-L expression on peripheral blood B cells. A significant increase in ICOS-L expression on resting CD38-CD19+ B cells in the Ambulatory cohort suggest that ICOS/ICOS-L mediated signaling is intact within Ambulatory donors (Fig. 2F). Hospitalized donors, however, display no ICOS-L expression on CD38-CD19+ B cells (Fig. 2F and Fig. S2). B cell activation and differentiation to antibody-secreting plasma cells is characterized by an increase in CD38 expression on CD19+ B cells. Similar flow profiling demonstrated a significant enhancement of ICOS-L in on CD38+ CD19+ B cells in Ambulatory donors (Fig. 2G). Hospitalized donors, however, display limited elevation of ICOS-L within the CD38+ CD19+ B cell fraction (Fig. 2G), suggesting impaired co-stimulation through the ICOS pathway and that B cell differentiation in severe CoViD-19, if present, may be driven through compensatory pathways that may result in sub-optimal B cell responses. Deficient ICOS expression and lack of functional ICOS/ICOS-L interactions in humans is associated with common variable immunodeficiency (CVID), which manifests as a decreased in memory class-switched B cells and poor germinal center formation ^26, 27^. Defective ICOS-L mediated co-stimulation may be a causal factor in inducing germinal center defects that have been recently reported in severe CoViD-19 ^15, 16^.

### In Vitro Stimulation Reveals Divergent Functional Response in COVID-19 Cohorts

As Tfh-B cell entanglement is driven by signaling in both directions, intrinsic defects in Tfh cells could mediate the lack of B cell response in severe disease. To assess the fundamental capability of peripheral blood Tfh cells to respond to TCR activation and co-stimulation, CD4 T cells from healthy donor, Ambulatory, and Hospitalized cohorts were activated with a standard combination of anti-CD3 and anti-CD28 antibodies. Following stimulation, the ability of Tfh cells to produce the effector cytokines IL-21 and IFNγ was assessed by flow cytometry. To account for observed decreases in the fraction of CD4 T cells in the peripheral blood of Hospitalized CoViD-19 patients, the data was examined two ways: both as polyfunctional cells as a percent of cTfh as well as polyfunctional cells as a percent of total leukocytes to account for lymphocyte changes between subjects and cohorts. Intracellular staining demonstrated a strong and significantly elevated polyfunctional cytokine response in the Ambulatory cohort, with IL-21+ IFNγ+ Tfh cells encompassing mean of 35% of total cTfh cells relative to 8% in the Hospitalized cohort, which represents a four-fold reduction (Fig. 3A). When assessed as a fraction of total leukocytes, polyfunctional cTfh cells were significantly elevated in the Ambulatory cohort and represented a mean of 2% of total leukocytes (Fig. 3B). In contrast, polyfunctional cTfh cells only accounted for a mean of 0.03% of total leukocytes in Hospitalized CoViD-19 subjects, representing an average of 67-fold reduction relative to the Ambulatory cohort. Relative to age-matched healthy donor subjects, no expansion of polyfunctional cTfh cells was observed in Hospitalized CoViD-19 subjects (Fig. 3B). Similar trends are observed in the frequencies of cTfh cell expressing single cytokines, and data are shown in Figure S4. These data reveal a diminished capacity to produce IL-21 alone or in combination with IFNγ within Tfh cells of Hospitalized donors. IL-21 is a key cytokine for Tfh-mediated B cell responses, and its expression has been demonstrated to be regulated by ICOS-driven increases in c-Maf expression. In Hospitalized CoViD-19 donors, the failure to upregulate ICOS-L expression on B cells and the corresponding lack of IL-21 production by Tfh cells may mechanistically contribute to the diminished humoral response associated with severe disease.

**Fig. 3:**
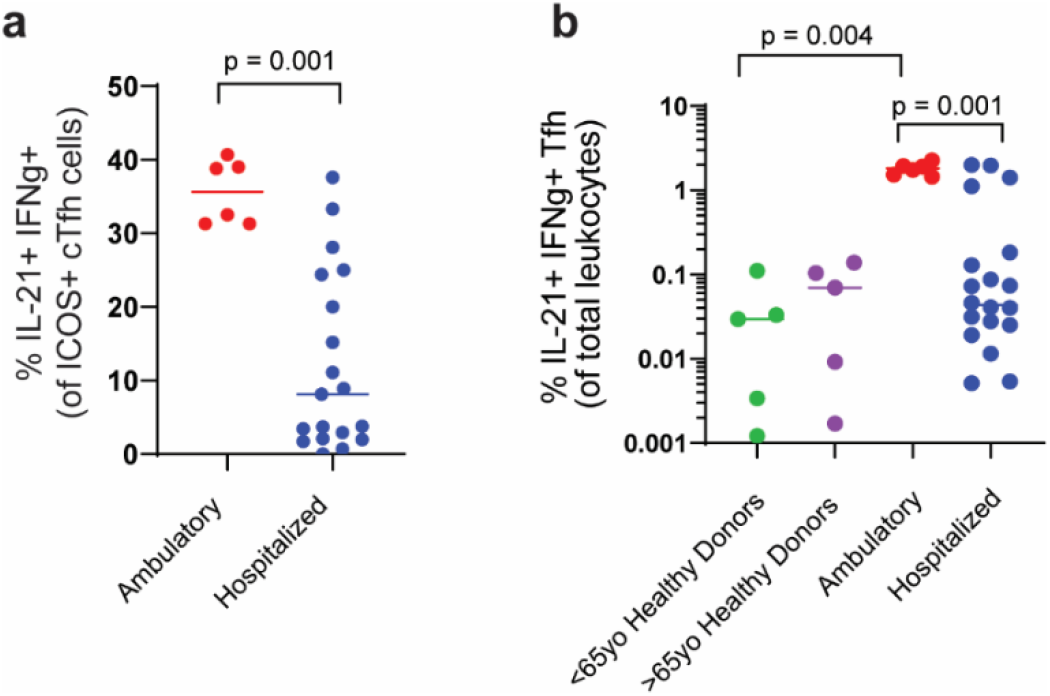
cTfh cells (ICOS+ CXCR5+ CD4+ cells) in Hospitalized CoViD-19 Donors are Defective in IL-21 and IFNγ Production. **a**, The detection of IL-21 and IFNγ producing ICOS+ cTfh cells following ex vivo anti-CD3/CD28 stimulation. Detection of cells producing both IL-21 and IFNγ by Ambulatory (n=6) and Hospitalized (n=20) donors represented as fraction of total ICOS+ Tfh cells. **b**, detection of ICOS+ cTfh cells producing both IL-21 and IFNγ in <65yo healthy donors (N=5), >65yo healthy donors (N=5), Ambulatory (N=6), and Hospitalized (N=20) donors represented as a fraction of total leukocytes. Means are shown (horizontal lines). Statistical significance was assessed by Kolmogorov-Smirnov test of group distributions.

### Co-Stimulation through ICOS Pathway Improves Defective Functional Response

To assess if exogenous ICOS co-stimulation may enhance cytokine production within Tfh cells of CoViD-19 donors, a parallel experiment as above was performed where peripheral blood mononuclear cells (PBMCs) were stimulated with a combination of sub-optimal anti-CD3 and saturating anti-CD28 or sub-optimal anti-CD3 and saturating anti-ICOS agonist antibody. The concentration of anti-CD3 used was sufficient to induce T cell activation but not induce proliferation or cytokine secretion on its own, providing a window to assess the effects of specific co-stimulation. After a 24hr incubation, expression of the effector cytokines IL-21 and IFNγ was assessed by flow cytometry. Similar to the previous experiment, robust cytokine responses were observed in the Ambulatory cohort, but a significant reduction in frequency of cells expressing both IL-21and IFNγ (Fig. 4A) was observed in the Hospitalized cohort with CD28 co-stimulation. In contrast to CD28, co-stimulation through ICOS resulted in a significant increase in the fraction of IL-21+ IFNγ+ Tfh cells (6-fold increase) in Hospitalized subjects (Fig. 4A).

**Fig. 4:**
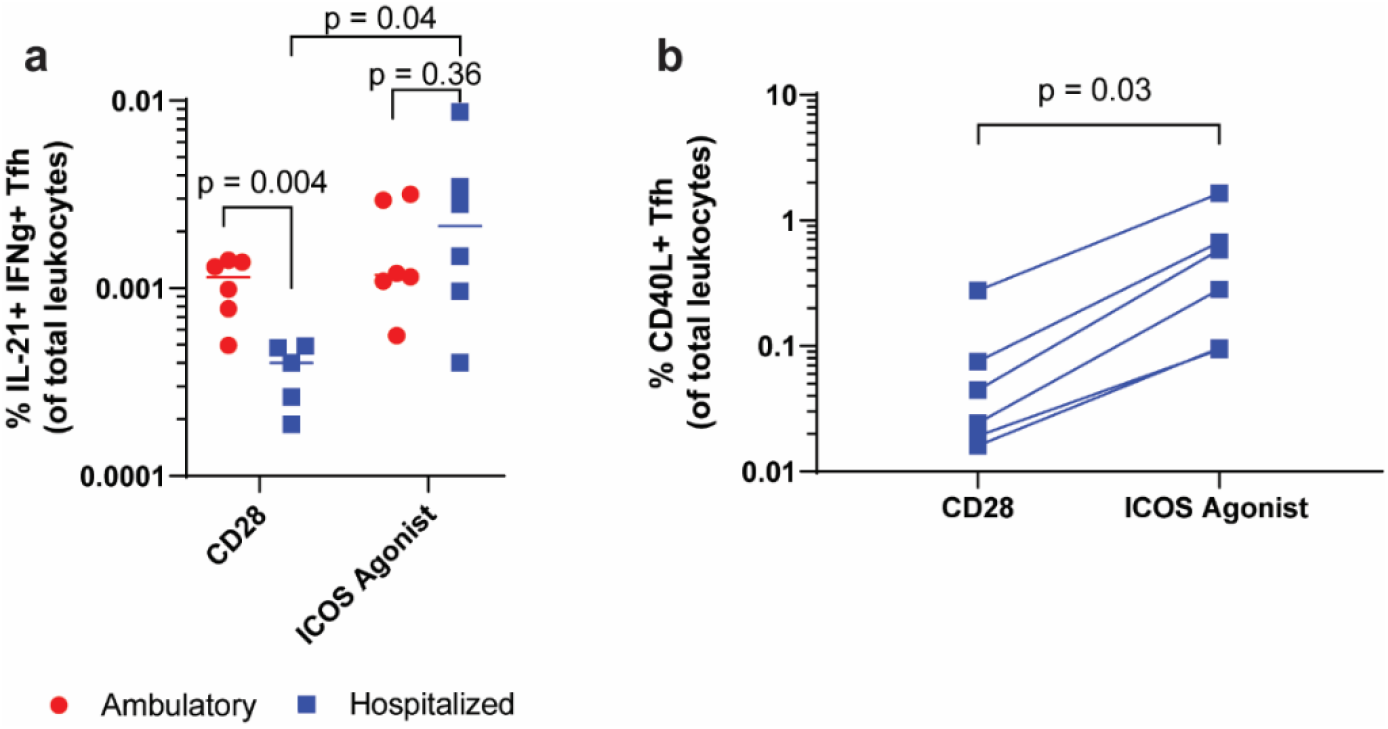
ICOS agonist antibody treatment increases polyfunctional cytokine response and increases CD40L expression by cTfh cells from Hospitalized CoViD-19 Donors. **a**, The detection of IL-21 and IFNγ producing ICOS+ cTfh cells following ex vivo anti-CD3/CD28 or CD3/ICOS agonist stimulation by Ambulatory (n=6) and Hospitalized (n=6) donors. Means are shown (horizontal lines). **b**, Assessment of frequency of CD40-L expressing Tfh cells following ex vivo stimulation with anti-CD3/CD28 or anti-CD3/ICOS agonist in samples from Hospitalized (N=6) donors. Statistics were assessed within subjects using a Wilcoxon Signed-Ranked test and across cohorts using Kolmogorov-Smirnov test of group distributions.

When examined across Ambulatory and Hospitalized cohorts, co-stimulation through ICOS resulted in polyfunctional cytokine expression by Tfh cells that was similar to the fraction observed in Ambulatory subjects as a trend towards elevated frequency of polyfunctional cTfh cells in hospitalized relative to ambulatory subjects was observed following ICOS agonism (Fig. 4A). Similar increases in cTfh cells producing a single-cytokine was also observed (Fig. S4C-D). ICOS signaling in Tfh cells has been reported to result in increased expression of CD40-L, which is essential for the stimulation of B cell differentiation and affinity maturation of B cell receptors ^22, 28^. ICOS agonism resulted in a significant increase of CD40-L expression (5-fold average increase) on anti-CD3 primed Tfh cells (Fig. 4B). Increasing of IL-21 production and CD40-L expression through ICOS agonism, as demonstrated following polyclonal T cell stimulation, suggests therapeutic interventions aimed at agonizing the ICOS receptor may modulate the Tfh-B cell axis and promote a functional humoral response in CoViD-19 patients.

## Discussion

While host immune responses aren’t necessarily altered upon viral infection, aberrant immune responses have been observationally described in CoViD-19 subjects, and the overall proficiency and durability of the adaptive immune response is poorly understood. Recent studies have demonstrated detection of robust T cell responses to antigenic SARS-CoV-2 proteins, but antibody responses have been observed to peak approximately 3-4 weeks post-symptom onset and then taper over the course of follow-up ^13, 29^. This delayed B cell response and the subsequent loss of humoral immunity despite a protracted course of disease in CoViD-19 subjects raises concerns over the natural robustness of long-term immunity to SARS-CoV-2 that may mirror waning immunity observed with SARS-CoV-1 ^30, 31^. Pathological studies in a cohort of deceased CoViD-19 subjects demonstrated a lack of T and B cell responses in lung tissue and a stark ablation of germinal centers within draining hilar lymph nodes ^15^. A separate study on lymph node and spleen biopsies demonstrated a similar pattern in live CoViD-19 subjects ^16^. The published observational data provide further support of the hypothesis that Tfh-mediated B cell reactions are abrogated in severe CoViD-19.

Mechanistically, Tfh cells appear to be the cornerstone for triggering and maintaining productive B cell responses. The data presented in this manuscript suggest that an impairment in Tfh-mediated B cell response may be contributing to the observed responses in severe CoViD-19 donors. Whether the lack of B-cell-mediated co-stimulation of Tfh cells or Tfh cell-mediated B cell activation is causal is yet unknown, but the two dysfunctional mechanisms are evident and may play a role in severe disease. ICOS-mediated co-stimulation of Tfh cells leads to increased secretion of IL-21 and CD40L expression, which are key mediators of B cell activation and plasmablast differentiation. Data presented herein suggest an intrinsic defect in Tfh-mediated cytokine expression due to failure to upregulate ICOS-L in Hospitalized CoViD-19 subjects. Providing additional activation of Tfh cells through treatment with an ICOS agonist antibody in hospitalized CoViD-19 subjects characterized by this defect, may act to stimulate the natural Tfh-B cell immunity. This may drive both increased kinetics of antibody responses, contribute to accelerated resolution of disease and facilitate more durable immunity. The observed ability of ICOS agonism, but not a traditional costimulatory molecule like CD28, to increase IL-21 responses demonstrates the uniqueness of ICOS-mediated co-stimulation in Tfh cell responses. ICOS agonist antibodies in preclinical development have been demonstrated to induce robust CD4 activity, including proliferation, activation, and cytokine secretion in vitro ^32^. This in vitro activity has carried over to the clinic, where treatment with an ICOS agonist in the setting of oncology demonstrated an ability to expand and maintain cTfh cells as well as Th1 and Tcm cells in the peripheral blood of long-term responders ^33^. Oncology studies provide precedent for testing a mechanism observed in the clinic that may be advantageous in boosting immune responses in the setting of hospitalized CoViD-19 subjects where natural B-cell mediated co-stimulation is deficient. The lack of co-stimulation may result not only in the absence of a necessary second signal for a durable immune response, but also exacerbate the lymphopenia as constant TCR stimulation in the absence of co-stimulation has been shown to result in T cell apoptosis ^34, 35^. Overall, the data suggest providing additional co-stimulation to the ICOS pathway may help restore and maintain a holistic humoral immune response, particularly in scenarios in which ICOS-L expression appears defective.

## Materials and Methods

### Study populations

Three cohorts were identified for the study.

The Ambulatory cohort was recruited through word of mouth from the population of front-line physicians and advance practice providers in Buffalo, NY as a part of a healthy volunteer study, JTX-HV-002. Eligible participants had a self-reported history of SARS-CoV-2 infection with positive PCR test who met the definition of recovered by the WHO criteria for healthcare workers. Study was approved through the New England IRB (Needham, MA). Participants provided three peripheral blood and serum samples over a 3 to 6-week period and information regarding severity and duration of symptoms was obtained at time of informed consent.

The Hospitalized cohort samples were obtained from Tissue Solutions (Glasgow, UK) through a collaboration with hospitals within the National Health Services network. Hospitalized patients who were found to have a positive COVID-19 PCR, or those with history, imaging, and clinical course strongly suggestive of COVID-19 despite negative testing were included in the cohort. Data from the medical record was abstracted by a third party (Tissue Solutions) and included date of positive test, medications, current treatment, previous symptoms, and hospital course through the point of data abstraction. Severity of disease was categorized in accordance with the WHO R&D Blueprint for the novel Coronavirus^36^.

The control cohort was selected from a pool of healthy donors obtained from Discovery Life Sciences (Huntsville, AL), with samples collected between 11/2016-11/2018, prior to the SARS-CoV-2 pandemic. Informed consent was obtained and documented for all subjects enrolled.

### Sample Processing

Peripheral blood samples from the Ambulatory and Hospitalized cohorts were collected directly into 8mL sodium heparin CPT tubes using a BD 70vacutainer safety-lok blood collection system and processed according to manufacturer’s instructions with the following modifications. Immediately after collection the CPT tube was gently inverted 10 times to mix the anticoagulant with the blood. CPT tubes were then spun at 1800g for 15 min. at 22°Cin a swing bucket centrifuge, cell and plasma fractions were then decanted into a 15mL Falcon tube and spun at 300g for 15 min. at 22°C. Supernatant was decanted, and the cell pellet was re-suspended in 1mL freezing media (10% DMSO in FBS) and transferred via pipet to a 1.8mL cryovial. Cryovials were immediately placed in a room temperature Mr. Frosty™ (Thermo Fisher Scientific) or CoolCell® (BioCision, LLC) and stored overnight in a − 80°C freezer and then shipped directly on packed in dry ice. Upon receipt, cryovials were immediately stored at − 80°C until flow cytometry analysis.

### Immunophenotyping of PBMC samples

PBMCs were counted and 1×10^6^ cells were reserved for staining with one of two flow panels to assess immunophenotypic populations of interest. In flow panel one, samples were re-suspended in flow buffer (PBS+2% FBS, 1 mM EDTA, and 0.1% Sodium azide) containing the following antibodies: CD3-PercpCy5.5 (clone OKT3, Biolegend), CD4-BUV805 (clone SK3, BD Biosciences), CXCR5-APC (clone J252D4, Biolegend), CXCR3-BUV395 (clone IC6/CXCR3, BD Biosciences), PD-1-BV421 (clone EH12.2H7, Biolegend) CD45RA-BUV737 (clone HI100, BD Biosciences), CD62L-PE-Cy7(clone DREG-56, Biolegend), CD19-BV785 (clone SJ25C1, Biolegend), CD38-BV605 (clone HB-7, Biolegend), ICOS-L-BUV661 (clone 2D3/B7H2, BD Biosciences) and eFluor780 fixable viability dye (eBioscience) for 30min at 4°C. After washing with flow buffer, cells were permeabilized and fixed using the FOXP3 Fixation/Permeabilization Concentrate and Diluent Kit (eBioscience) according to the manufacturer’s instructions. Permeabilized cells were stained with biotinylated anti-human ICOS antibody (clone M13, Jounce Therapeutics) and BCL2-AF488 (clone 100, Biolegend) for 30 min at RT. After washing, cells were incubated with streptavidin-PE (Biolegend) for 30 min at RT. Cells were then washed and re-suspended in flow buffer. Flow cytometry was performed using the LSR Fortessa (BD Biosciences).

In flow panel two, samples were re-suspended in flow buffer (PBS+2% FBS, 1 mM EDTA, and 0.1% Sodium azide) containing the following antibodies: CD3-PercpCy5.5 (clone OKT3, Biolegend), CD4-BUV805 (clone SK3, BD Biosciences), CXCR5-APC (clone J252D4, Biolegend), CD19-BV785 (clone SJ25C1, Biolegend), CD38-BUV395 (HB-7, BD Biosciences), HLA-DR-BV510 (clone G46-6, BD Biosciences), CD154-PeCy7 (clone 24-31, Biolegend), CD27-FITC (clone: M-T271, BioLegend) and eFluor780 fixable viability dye (eBioscience) for 30min at 4°C. After washing with flow buffer, cells were permeabilized and fixed using the FOXP3 Fixation/Permeabilization Concentrate and Diluent Kit (eBioscience) according to the manufacturer’s instructions. Permeabilized cells were stained with biotinylated anti-human ICOS antibody (clone: M13, Jounce Therapeutics) for 30min at RT. Cells were washed and then stained with streptavidin-BV421 for 30min, RT. After washing, cells were re-suspended in flow buffer and flow cytometry was performed using the LSR Fortessa (BD).

### CD3/CD28 and CD3/anti-ICOS 36E10 Plate Bound Stimulation + Normal Donor and COVID Cohort CD3/CD28 Bead Stimulation

Plate bound antibody conditions were prepared as follows: In 1X PBS in a 96 well round bottom TC plate, plate bound anti-human CD3 (clone: OKT3, BioXCell) was coated at a final sub-optimal stimulation concentration of 1ug/ml with either plate bound anti-human CD28 (clone:CD28.2, Biolegend) at a final concentration of 10ug/ml, or 10ug/ml final concentration of anti-human ICOS (clone: 36E10, Jounce Therapeutics) for 2 hours at 37C. Plates were washed with 1X PBS to remove un-bound antibody and cells were plated at .25×10^6 cells/ well, in triplicate, from ambulatory COVID subjects, or hospitalized COVID subjects. At this time, age representative younger and older normal donor (pre-COVID collection) PBMCs were also plated at .25×10^6 cells/ well, in triplicate in untreated wells of a 96 round bottom TC plate in the presence of CD3/CD28 beads (Gibco) at a 3:1 bead to T cell ratio, assuming ∼60% T cell composition within each well. Beads were prepared for plating according to manufacturer’s instructions. All plates were then incubated for 24hr at 37C. Brefeldin A (Biolegend) and monensin (Biolegend) were added according to manufacturer’s instructions for the final five to six hours of the incubation. Cells were then harvested and replicate wells across conditions and donors were combined. After combination, cells were transferred to a new 96 well round bottom plate and re-suspended in flow buffer (PBS + 2% FBS, 1 mM EDTA, and 0.1% Sodium azide) containing anti-human Fc Block for 20 min at RT. Cells were then washed with flow buffer and resuspended in a master extracellular staining mix containing the following antibodies: CD3-PerCpCy5.5 (clone: OKT3, Biolegend), CD4-BUV805 (clone SK3, BD Biosciences), CXCR5-APC (clone:J252D4, Biolegend) CD154-PECy7 (clone: 24-31 Biosciences), CD38-BUV395 (clone: HB7, BD Biosciences) and eFluor780 fixable viability dye (eBioscience) for 30 min at 4°C. After washing with flow buffer, cells were permeabilized and fixed using the FOXP3 Fixation/Permeabilization Concentrate and Diluent Kit (eBioscience) according to the manufacturer’s instructions. Permeabilized cells were stained with biotinylated anti-human ICOS antibody (clone: M13, Jounce Therapeutics), IL-21-PE (clone: 3A3-N2, Biolegend), and IFNg-BV605 (clone:B27, Biolegend) for 30 min, RT. Cells were then washed and stained with streptavidin-BV421 for 30min, RT. After washing, cells were re-suspended in flow buffer and flow cytometry was performed using the LSR Fortessa (BD). Samples acquisition was performed by transferring cells from the 96 well round bottom staining plate to a 5ml FACS tube and running each sample dry.

In a separate experimental preparation, ambulatory and hospitalized COVID subject samples were plated at .25×10^6 cells/ well in singlet in untreated wells of a 96 round bottom TC plate in the presence of CD3/CD28 beads (Gibco) at a 3:1 bead to T cell ratio, assuming ∼60% T cell composition within each well. Beads were prepared for plating according to manufacturer’s instructions and were then incubated for 24hr at 37C. Brefeldin A (Biolegend) and monensin (Biolegend) were added according to manufacturer’s instructions for the final five to six hours of the incubation. Cells were then harvested and transferred to a new 96 well round bottom plate and re-suspended in flow buffer (PBS + 2% FBS, 1 mM EDTA, and 0.1% Sodium azide) containing anti-human Fc Block for 20 min at RT. Cells were then washed with flow buffer and resuspended in a master extracellular staining mix containing the following antibodies: CD3-PerCpCy5.5 (clone: OKT3, Biolegend), CD4-BUV805 (clone SK3, BD Biosciences), CXCR5-APC (clone:J252D4, Biolegend) CD154-PECy7 (clone: 24-31 Biosciences), CD38-BUV395 (clone: HB7, BD Biosciences) and eFluor780 fixable viability dye (eBioscience) for 30 min at 4°C. After washing with flow buffer, cells were permeabilized and fixed using the FOXP3 Fixation/Permeabilization Concentrate and Diluent Kit (eBioscience) according to the manufacturer’s instructions. Permeabilized cells were stained with biotinylated anti-human ICOS antibody (clone: M13, Jounce Therapeutics), IL-21-PE (clone: 3A3-N2, Biolegend), and IFNg-BV605 (clone:B27, Biolegend) for 30 min, RT. Cells were then washed and stained with streptavidin-BV421 for 30min, RT. After washing, cells were re-suspended in flow buffer and flow cytometry was performed using the LSR Fortessa (BD). Samples acquisition was performed by transferring cells from the 96 well round bottom staining plate to a 5ml FACS tube and running each sample dry.

### Statistical Analyses

All statistical analyses were performed in Prism (GraphPad Software). Comparison of the distribution of frequencies of cell phenotypes between cohorts was performed using the Kolmogorov-Smirnov test. Assessment of stimulation-induced changes were performed using a Wilcoxon Signed-rank test. A p value of < 0.05 was deemed significant. Calculated p values are indicated in graphs where the difference was statistically significant.

## Data Availability

All data necessary to support the conclusions of the manuscript are contained within the manuscript or supplemental materials.

## Acknowledgments

We would like to thank all the donors for use of their samples in this study. We would also like to thank Ashley Graca and Beth Trehu at Jounce Therapeutics for their assistance in the conduct of JTX-HV-002 and enrollment of the Ambulatory cohort.

## Funding

This work was sponsored and funded by Jounce Therapeutics, Inc.

## Author Contributions

A.H., H.C., A.D., H.W., C.S., and N.S. contributed to the design of experiments, experimental conduct, data acquisition and analysis, and writing of this manuscript. B.W.H. contributed to the conceptualization of the study, data analysis and interpretation, sample collection, and manuscript writing. R.M. and C.J.H. contributed to the conceptualization of the study, experimental design, data interpretation, and manuscript writing.

## Competing Interests

All authors are employees or contractors of Jounce Therapeutics, Inc.

## Notes

### Author Declarations

The study was approved through the New England IRB (Needham, MA).

## References

1. Johns Hopkins University. CoViD-19 Dashboard. (ed^(eds). Johns Hopkins University Center for Systems Science and Engineering.

2. Chen G, et al. Clinical and immunological features of severe and moderate coronavirus disease 2019. J Clin Invest 130, 2620–2629 (2020).

3. Chen Z, John Wherry E. T cell responses in patients with COVID-19. Nat Rev Immunol, (2020).

4. De Biasi S, et al. Marked T cell activation, senescence, exhaustion and skewing towards TH17 in patients with COVID-19 pneumonia. Nat Commun 11, 3434 (2020).

5. Kuri-Cervantes L, et al. Immunologic perturbations in severe COVID-19/SARS-CoV-2 infection. bioRxiv, (2020).

6. Le Bert N, et al. SARS-CoV-2-specific T cell immunity in cases of COVID-19 and SARS, and uninfected controls. Nature, (2020).

7. Mathew D, et al. Deep immune profiling of COVID-19 patients reveals patient heterogeneity and distinct immunotypes with implications for therapeutic interventions. bioRxiv, (2020).

8. Sekine T, et al. Robust T cell immunity in convalescent individuals with asymptomatic or mold COVID-19. bioRxiv, (2020).

9. Thevarajan I, et al. Breadth of concomitant immune responses prior to patient recovery: a case report of non-severe COVID-19. Nat Med 26, 453–455 (2020).

10. Fox A, et al. Severe pandemic H1N1 2009 infection is associated with transient NK and T deficiency and aberrant CD8 responses. PLoS One 7, e31535 (2012).

11. Tan L, et al. Lymphopenia predicts disease severity of COVID-19: a descriptive and predictive study. Signal Transduct Target Ther 5, 33 (2020).

12. Atyeo C, et al. Distinct early serological signatures track with SARS-CoV-2 survival. Immunty, (2020).

13. Wang Y, et al. Kinetics of viral load and antibody response in relation to COVID-19 severity. J Clin Invest, (2020).

14. Zohar T, Alter G. Dissecting antibody-mediated protection against SARS-CoV-2. Nat Rev Immunol 20, 392–394 (2020).

15. Duan YQ, et al. Deficiency of Tfh Cells and Germinal Center in Deceased COVID-19 Patients. Curr Med Sci, (2020).

16. Kaneko N, et al. The Loss of Bcl-6 Expressing T Follicular Helper Cells and the Absence of Germinal Centers in COVID-19. SSRN, 3652322 (2020).

17. Bauquet AT, et al. The costimulatory molecule ICOS regulates the expression of c-Maf and IL-21 in the development of follicular T helper cells and TH-17 cells. Nat Immunol 10, 167–175 (2009).

18. Bentebibel SE, et al. Induction of ICOS+CXCR3+CXCR5+ TH cells correlates with antibody responses to influenza vaccination. Sci Transl Med 5, 176ra132 (2013).

19. Bossaller L, et al. ICOS deficiency is associated with a severe reduction of CXCR5+CD4 germinal center Th cells. J Immunol 177, 4927–4932 (2006).

20. Kopf M, et al. Inducible costimulator protein (ICOS) controls T helper cell subset polarization after virus and parasite infection. J Exp Med 192, 53–61 (2000).

21. Liu D, et al. T-B-cell entanglement and ICOSL-driven feed-forward regulation of germinal centre reaction. Nature 517, 214–218 (2015).

22. McAdam AJ, et al. ICOS is critical for CD40-mediated antibody class switching. Nature 409, 102–105 (2001).

23. Roussel L, et al. Loss of human ICOSL results in combined immunodeficiency. J Exp Med 215, 3151–3164 (2018).

24. Kundig TM, et al. Duration of TCR stimulation determines costimulatory requirement of T cells. Immunity 5, 41–52 (1996).

25. Kuchen S, et al. Essential role of IL-21 in B cell activation, expansion, and plasma cell generation during CD4+ T cell-B cell collaboration. J Immunol 179, 5886–5896 (2007).

26. Salzer U, et al. ICOS deficiency in patients with common variable immunodeficiency. Clin Immunol 113, 234–240 (2004).

27. Warnatz K, et al. Human ICOS deficiency abrogates the germinal center reaction and provides a monogenic model for common variable immunodeficiency. Blood 107, 3045–3052 (2006).

28. Koike T, Harada K, Horiuchi S, Kitamura D. The quantity of CD40 signaling determines the differentiation of B cells into functionally distinct memory cell subsets. Elife 8, (2019).

29. Sun B, et al. Kinetics of SARS-CoV-2 specific IgM and IgG responses in COVID-19 patients. Emerg Microbes Infect 9, 940–948 (2020).

30. Liu A, et al. Disappearance of antibodies to SARS-CoV-2 in a -COVID-19 patient after recovery. Clin Microbiol Infect, (2020).

31. Tang F, et al. Lack of peripheral memory B cell responses in recovered patients with severe acute respiratory syndrome: a six-year follow-up study. J Immunol 186, 7264–7268 (2011).

32. Hanson A, et al. ICOS agonism by JTX-2011 (vopratelimab) requires initial T cell priming and Fc cross-linking for optimal T cell activation and anti-tumor immunity in preclinical models. PLoS ONE 15, (2020).

33. Hanson A, et al. Abstract 5536: ICOS hi CD4 T cells emerging on vopratelimab treatment have Th1, Tcm, and Tfh characteristics and may contribute to durability of clinical responses. Cancer Research 80, (2020).

34. Schwartz RH. A cell culture model for T lymphocyte clonal anergy. Science 248, 1349–1356 (1990).

35. Yamamoto T, Hattori M, Yoshida T. Induction of T-cell activation or anergy determined by the combination of intensity and duration of T-cell receptor stimulation, and sequential induction in an individual cell. Immunology 121, 383–391 (2007).

36. Organization WH. WHO R&D Blueprint novel Coronavirus COVID-19 Therapeutic Trial Synopsis. R&D Blueprint, (2020).

