## Supplemental Materials for "Impaired ICOS signaling between Tfh and B cells distinguishes hospitalized from ambulatory CoViD-19 patients"

### Supplementary Materials

Description of Study Cohorts

Fig.S1. Timepoint of sample collection relative to diagnosis of CoViD-19 for Ambulatory and Hospitalized subjects

Fig.S2. Trend toward an in ICOS-L expression on a per-cell basis is only observed in Ambulatory but not Hospitalized CoViD-19 donors

Fig.S3. Tfh cell frequencies in <65 and >65-year-old healthy donor subjects.

Fig.S4. Frequencies of single cytokine producing cTfh cells.

Fig.S5**.** Gating strategy for assessment of cTfh frequencies and representative examples.

Fig.S6. Gating strategy for Tfh specific cytokine response with CD3 and CD28 or ICOS agonist treatment.

Table S1. Characterization of Study Cohorts.

**Description of Study Cohorts**

By utilizing the WHO R&D Blueprint for the novel Coronavirus, severity of illness was scored for each participant, and the distribution is described in Table S1. Healthy donor (HD) controls were considered uninfected (no clinical or virological evidence of infection, Score 0). The Ambulatory cohort scored as a 1 or 2 (no limitation of activities (15%), limitation of activities (85%) respectively), whereas the Hospitalized cohort was scored 3-8. Hospitalization was further broken down into Mild disease (score 3 no oxygen therapy (35%), score 4 oxygen via face mask or nasal cannula (35%)), Severe (score 5 non-invasive ventilation or high flow oxygen (0%), score 6 intubation and mechanical ventilation (5%), score 7 ventilation and additional support (15%)) and Death (score of 8 (10%)). The control cohort samples were collected between 11/2016-11/2018, prior to the SARS-CoV-2 pandemic.

Participants in the Ambulatory cohort had a median age of 29 (27-44) and were the youngest cohort, without any individuals over the age of 50. In contrast, the Hospitalized cohort was significantly older, with a median age of 74.5 (36-95) and healthy donors (phenotyping cohort) had a median age of 37 (28-84). There was a female predominance in the Ambulatory and Healthy donor (phenotyping cohort) groups (61.5% and 57% respectively), compared to the Hospitalized cohort (30% female). All cohorts were predominantly white, non-Hispanic. The median length of time from positive test to first sample collection was 35 (10-64) days for the Ambulatory cohort and 21 (2-61) for the Hospitalized cohort (Fig. S1).

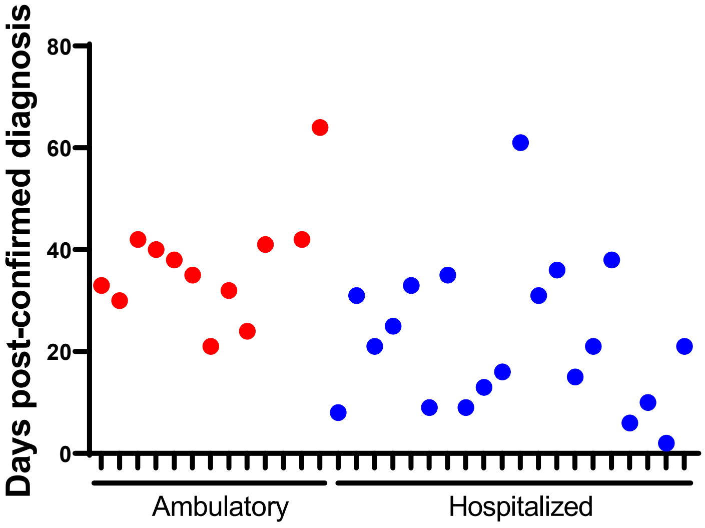


**Fig. S1.**

Timepoint of sample collection relative to diagnosis of CoViD-19 for Ambulatory and Hospitalized subjects. Time in days from CoViD-19 diagnosis to collection of PBMC samples was determined for subjects in both Ambulatory and Hospitalized cohorts.

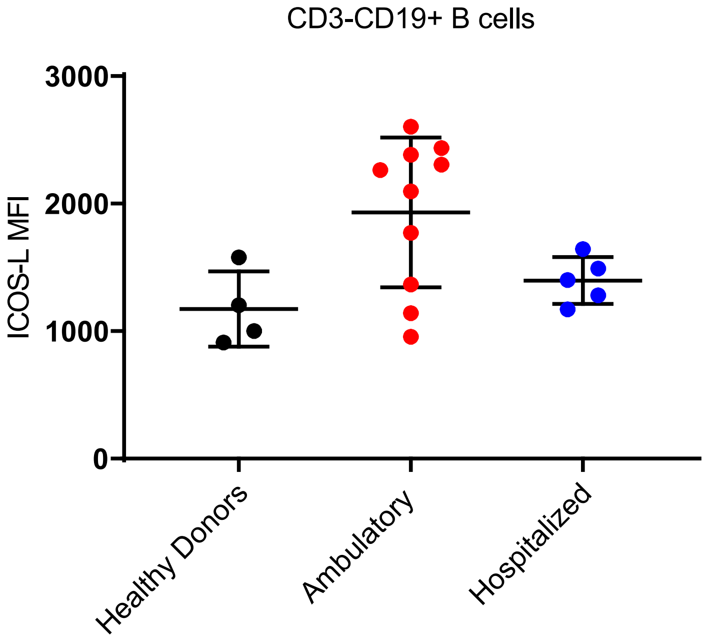


**Fig. S2.**

Trend toward an in ICOS-L expression on a per-cell basis is only observed in Ambulatory but not Hospitalized CoViD-19 donors. The expression of ICOS-L by CD19+ B cells on a per cell basis was assessed by measuring fluorescence intensity by flow cytometry.

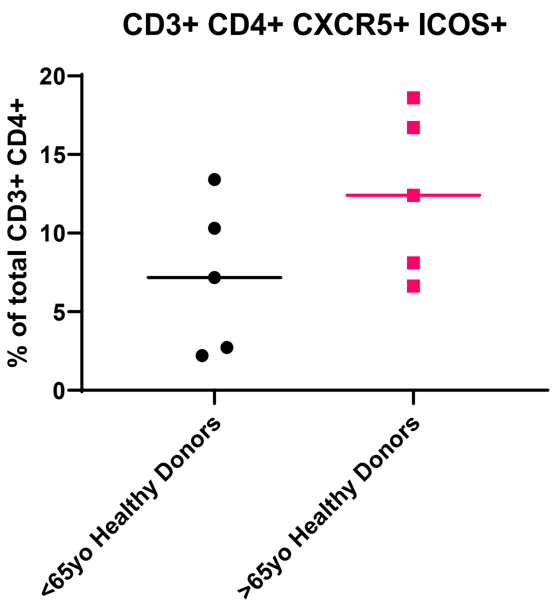


**Fig. S3.**

Tfh cell frequencies in <65 and >65-year-old healthy donor subjects. The frequency of by ICOS+ CXCR5+ Tfh cells was assessed by measuring fluorescence intensity by flow cytometry.

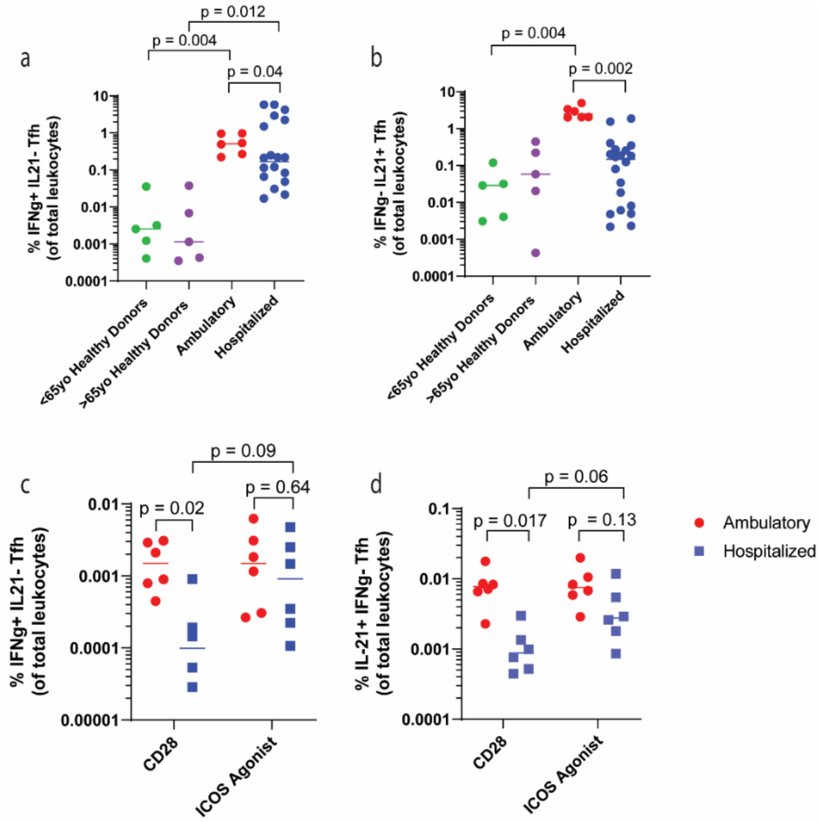


**Fig. S4.**

Frequencies of single cytokine producing cTfh cells. Frequency of single cytokine producing cTfh cells was assessed by flow cytometry. a, frequency of IFNg producing cells and b, IL-21 producing cells in <65-year-old healthy donors (n=6), >65 year old healthy donors (N=6), ambulatory (N=6), or hospitalized CoViD-19 (N=20) subjects following stimulation with CD3/CD28 beads. c, frequency of IFNg producing cTfh cells and d, IL-21 producing cTfh cells following either anti-CD3 plus anti-CD28 or anti-CD3 plus anti-ICOS agonist stimulation of ambulatory (N=6) or hospitalized (N=20) CoViD-19 subjects. Statistics were assessed within subjects using a Wilcoxon Signed-Ranked test and across cohorts using Kolmogorov-Smirnov test of group distributions.

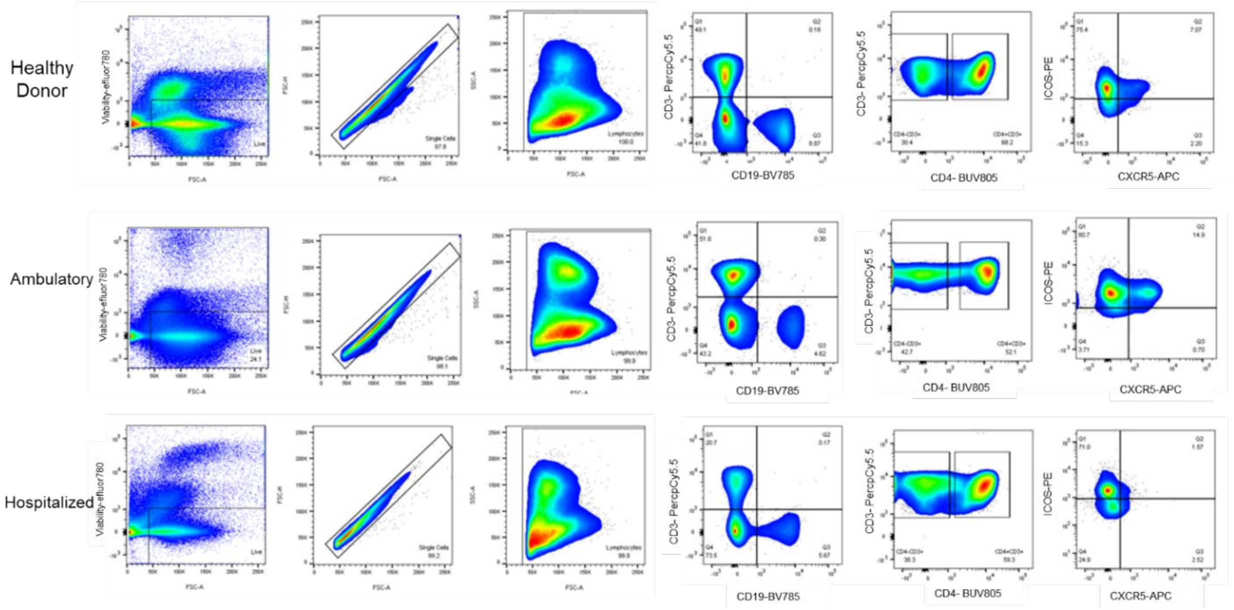


**Fig. S5.**

Gating strategy for assessment of cTfh frequencies and representative examples.

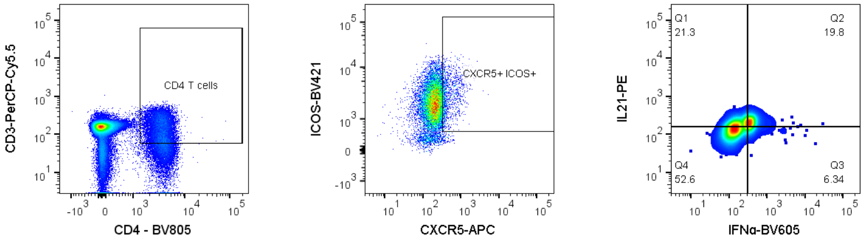


**Fig. S6.**

Gating strategy for Tfh specific cytokine response with CD3 and CD28 or ICOS agonist treatment.

| | Healthy Donors | | | Ambulatory (n=13) | Hospitalized (n=20) |
|---|---|---|---|---|---|
| | Phenotyping (N=7) | <65 years old (N=5) | >65 years old (N=5) | | |
| Age, Median (Range) | 37 (28-84) | 34 (28-40) | 79 (70-89) | 29 (27-44) | 74.5 (36-95) |
| Gender | | | | | |
| Male | 3 (34%) | 3 (60%) | 3 (6%) | 5 (38.5%) | 14 (60%) |
| Female | 4 (57%) | 2 (40%) | 2 (40%) | 8 (61.5%) | 6 (40%) |
| Race | | | | | |
| Caucasian | 7 (100%) | 5 (100% | 5 (100%) | 8 (61.5%) | 12 (60%) |
| Black | 0 | 0 | 0 | 1 (8.5%) | 0 |
| Asian | 0 | 0 | 0 | 2 (15%) | 1 (5%) |
| Other | 0 | 0 | 0 | 2 (15%) | 7 (35%) |
| Severity | | | | | |
| Uninfected, 0 | 7 (100%) | 5 (100%) | 5 (100%) | 0 | 0 |
| Ambulatory, 1 | 0 | 0 | 0 | 2 (15%) | 0 |
| 2 | 0 | 0 | 0 | 11 (85%) | 0 |
| Hospitalized, Mild, 3 | 0 | 0 | 0 | 0 | 7 (35%) |
| 4 | 0 | 0 | 0 | 0 | 7 (35%) |
| Hospitalized, Severe, 5 | 0 | 0 | 0 | 0 | 0 |
| 6 | 0 | 0 | 0 | 0 | 1 (5%) |
| 7 | 0 | 0 | 0 | 0 | 3 (15%) |
| Death, 8 | 0 | 0 | 0 | 0 | 2 (20%) |

**Table S1.**

Characterization of Study Cohorts. Demographics and disposition of subjects from whom samples were profiled for phenotype or biological activity. Three healthy donor cohorts of samples were used: one for flow-based phenotyping and two were age-matched to the Ambulatory or Hospitalized CoViD-19 cohorts. CoViD-19 severity was scored according to the WHO R&D Blueprint for the novel Coronavirus.
